# SARS-CoV-2 neutralizing antibodies in Chile after a vaccination campaign with five different schemes

**DOI:** 10.1101/2022.05.03.22274622

**Authors:** Ximena Aguilera, Juan Hormazabal, Cecilia Vial, Lina Jimena Cortes, Claudia Gonzalez, Paola Rubilar, Mauricio Apablaza, Muriel Ramirez-Santana, Gloria Icaza, Loreto Nuñez-Franz, Carla Castillo-Laborde, Carolina Ramirez-Riffo, Claudia Perez, Ruben Quezada-Gate, Macarena Said, Pablo Vial

**Affiliations:** Universidad del Desarrollo, Santiago, Chile; Universidad Católica del Norte, Coquimbo, Chile; Universidad de Talca, Talca, Chile; Clínica Alemana de Santiago, Chile

**Keywords:** BNT162b2, CoronaVac, AZD1222, COVID-19, SARS-CoV-2, Neutralizing antibodies, Chile, Santiago, Talca, Coquimbo, La Serena, coronavirus disease, general population, cross sectional design, respiratory infections, severe acute respiratory syndrome coronavirus 2, vaccine-preventable diseases, viruses

## Abstract

Using levels of neutralizing antibodies (nAbs), we evaluate the successful Chilean SARS-CoV-2 vaccine campaign, which combines technologies and heterologous boosters. In 120 randomly selected seropositive individuals from a population-based study, we conclude that the booster dose, regardless of vaccine technology or natural infection, and mRNA vaccines significantly improve nAbs response.

## Introduction

The SARS-CoV-2 pandemic has unprecedented challenges for its global, regional, and national control. The continuous emergence of the SARS-CoV-2 variants, jointly with the waning antibody titers from natural and vaccine-induced immunity, generates scenarios that maintain population susceptibility and risk of outbreaks (1,2). Chile is not the exception, presenting one of the worst outbreaks in the world by mid-2020, but also with a globally successful vaccine campaign. The Chilean vaccination strategy combined different vaccine technologies and heterologous boosters (3). We aim to compare the various vaccination schemes, using the presence of neutralizing antibodies (nAbs) as a correlate of immune protection against SARS-CoV-2 (4,5).

The Ethics Committees of the Universities el Desarrollo and Talca and the Facultad de Medicina of the Universidad Católica del Norte approved the study protocols. Informed consent was obtained from all subjects, if subjects are under 18, from a parent or legal guardian.

### The study

Serum neutralization capacity was measured using a pseudotyped vesicular stomatitis virus with a sequence encoding the enhanced green fluorescent protein as a reporter gene (VSV-GFP-Spike SARS-CoV-2 original Wuhan strain) kindly donated by Dr. Kartik Chandran (6). Samples tested came from individuals enrolled in a population-based SARS-CoV-2 seroprevalence study performed by the same research team (7–9). In November 2021, we collected 2,198 serum samples from seven-year-old and older people, finding 97.3% of seropositivity. We used six groups of positive samples according to natural infection history and the five most frequent vaccination schemes, selecting randomly 20 individuals from each group (Table 1).

**Table 1.** Vaccination schemes and sample distribution, Chile 2021.

| Vaccine scheme acronym | Description | N |
| --- | --- | --- |
| CC (CoronaVac CoronaVac) | Basal scheme = two doses of Sinovac's CoronaVac | 20 |
| PP (Pfizer Pfizer) | Basal scheme = two doses of BNT162b2 | 20 |
| CCO (CC plus Oxford AstraZeneca) | Basal CC plus booster with ChAdOx1-S (heterologous) | 20 |
| CCP (CC plus Pfizer) | Basal CC plus booster with BNT162b2 (heterologous) | 20 |
| PPP (Triple Pfizer) | Basal PP plus booster with BNT162b2 | 20 |
| Natural infection | Non-vaccinated, but seropositive (Natural Infection) | 20 |

The amount of nAbs response was measured as the inhibitory concentration where 50% of the viral entrance is inhibited (IC50). IC50 was calculated for each serum by measuring the viral entrance of the VSV-GFP-Spike SARS-CoV-2 pseudotype capturing the amount of GFP fluorescence in each serum dilution. Briefly, serum serial dilutions from 1/50 to 1/51200 were incubated with VSV-GFP-Spike SARS-CoV-2 pseudovirus for 30 minutes, and then VEROE6 cells (ATCC) were infected with this virus. After 20 hours, cells were washed, fixed in 4% paraformaldehyde, and GFP intensity was measured in a Cytation 3 (BioTeK). The resulting curve of each serum was analyzed through a dose-response nonlinear regression in Prism v9 Software (Graphpad) to calculate the IC50.

We found nAbs response in 82.5% of the subjects, without significant differences by sex or age. The presence of nAbs is significantly higher in people with booster doses and non-smokers. Also, it varies according to vaccine platform used (inactivated, mRNA or viral vector recombinant) (Table 2).

**Table 2.**
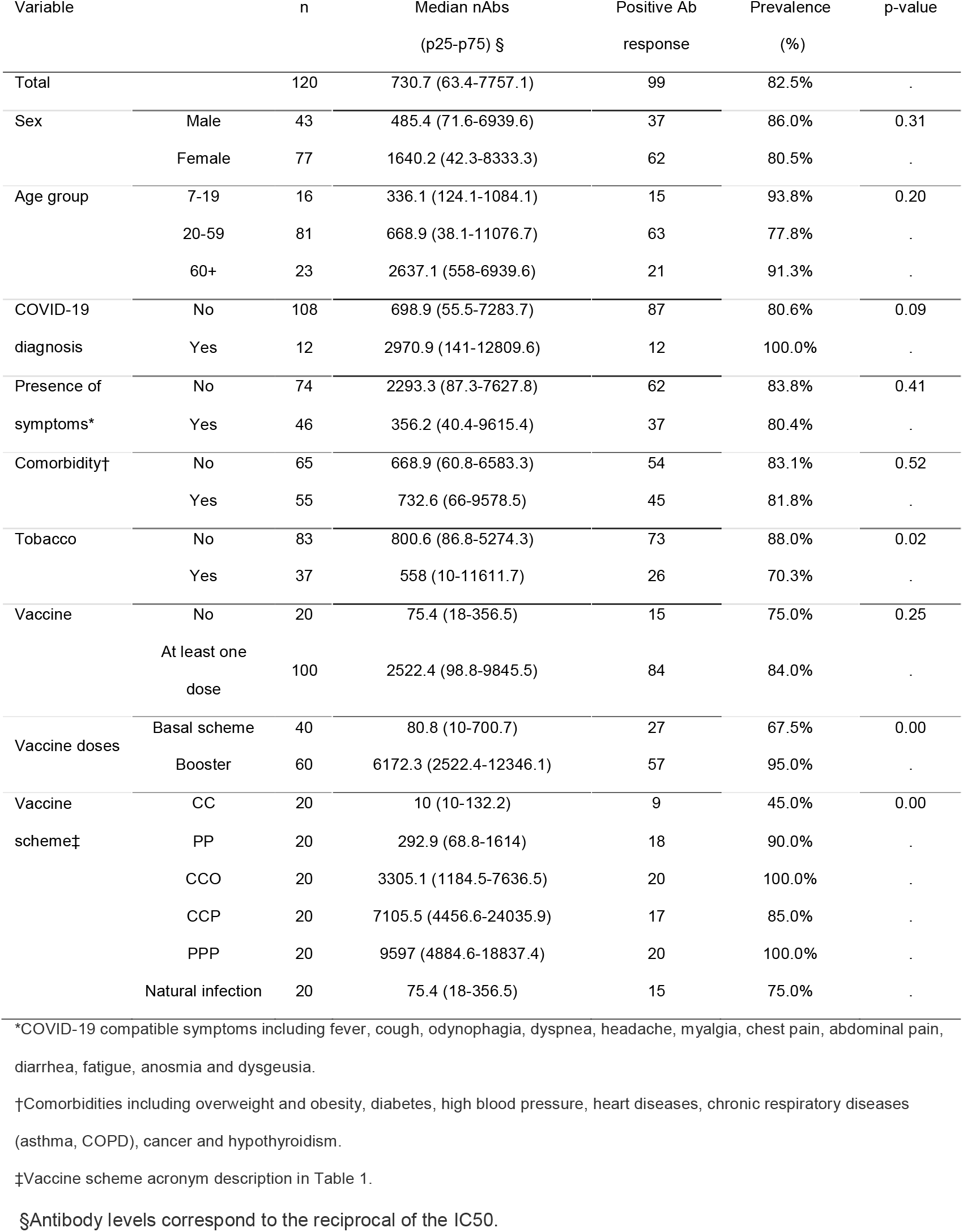
Presence of Neutralizing Antibodies against SARS-CoV-2 among seropositive individuals according to selected variables, Chile November 2021.

Figure 1 shows the level of neutralizing antibodies represented as median and interquartile values of the IC50 for each study group. In the left panel, when comparing nAbs levels, the group with only a basal immunization scheme has nAbs levels similar to that of the naturally infected patients (p value=0.8425). In contrast, individuals who received a booster dose have a significantly higher level of nAbs compared to the other two groups.

**Figure 1:**
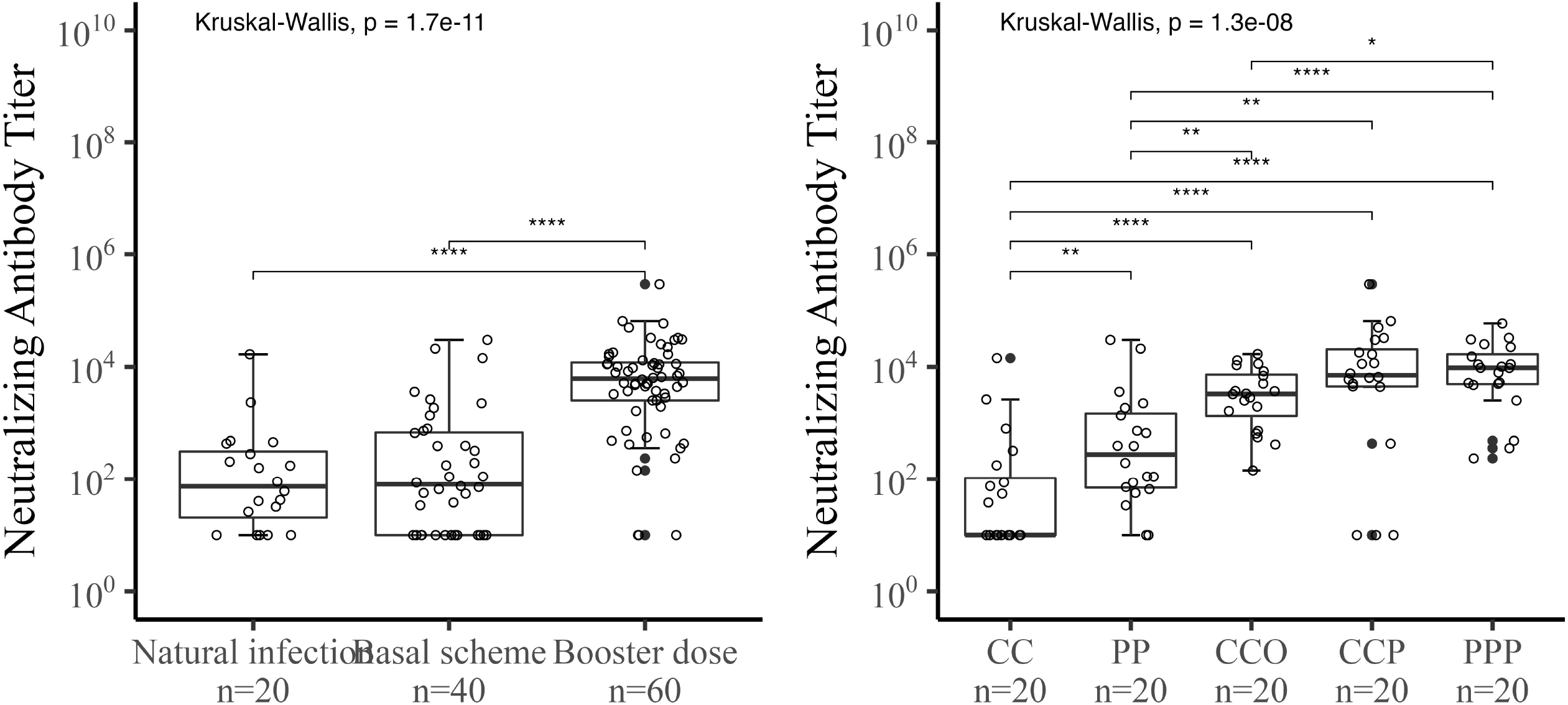
All samples were plotted as individuals points and graphed the median and interquartile for both panels. Left panel shows a comparison of nAbs titers between natural infection (n=20), two dose schemes (n=40), and booster dose (n=60). The right panel shows a comparison of nABs between the different vaccination schemes: CC (CoronaVac CoronaVac), PP (Pfizer Pfizer), CCO (CC plus Oxford AstraZeneca), CCP (CC plus Pfizer), PPP (Triple Pfizer), with n=20 for each group. The statistical differences were performed with kruskal-Wallis test, and a p-value < 0.05.

On the right panel of Figure 1, analyzing the schemes by the different vaccines used, it is observed that the PPP scheme elicited the highest median nAbs response, without significant differences with the heterologous CCP scheme, but higher than the CCO scheme. On the other hand, all three booster schemes produced significantly higher nAbs levels than the natural infection group and the two basal schemes studied (CC and PP). Among the basal schemes, there are also significantly higher nAbs levels for the scheme with mRNA vaccines (PP) compared to inactivated vaccines (CC).

Figure 2 is a scatter plot showing the relationship between nAbs titers and time, using days since the last vaccine dose. It shows the waning of antibody titers for the groups with the basal vaccine scheme, but not for the groups with the booster doses, but also the follow-up was shorter for the latter groups.

**Figure 2:**
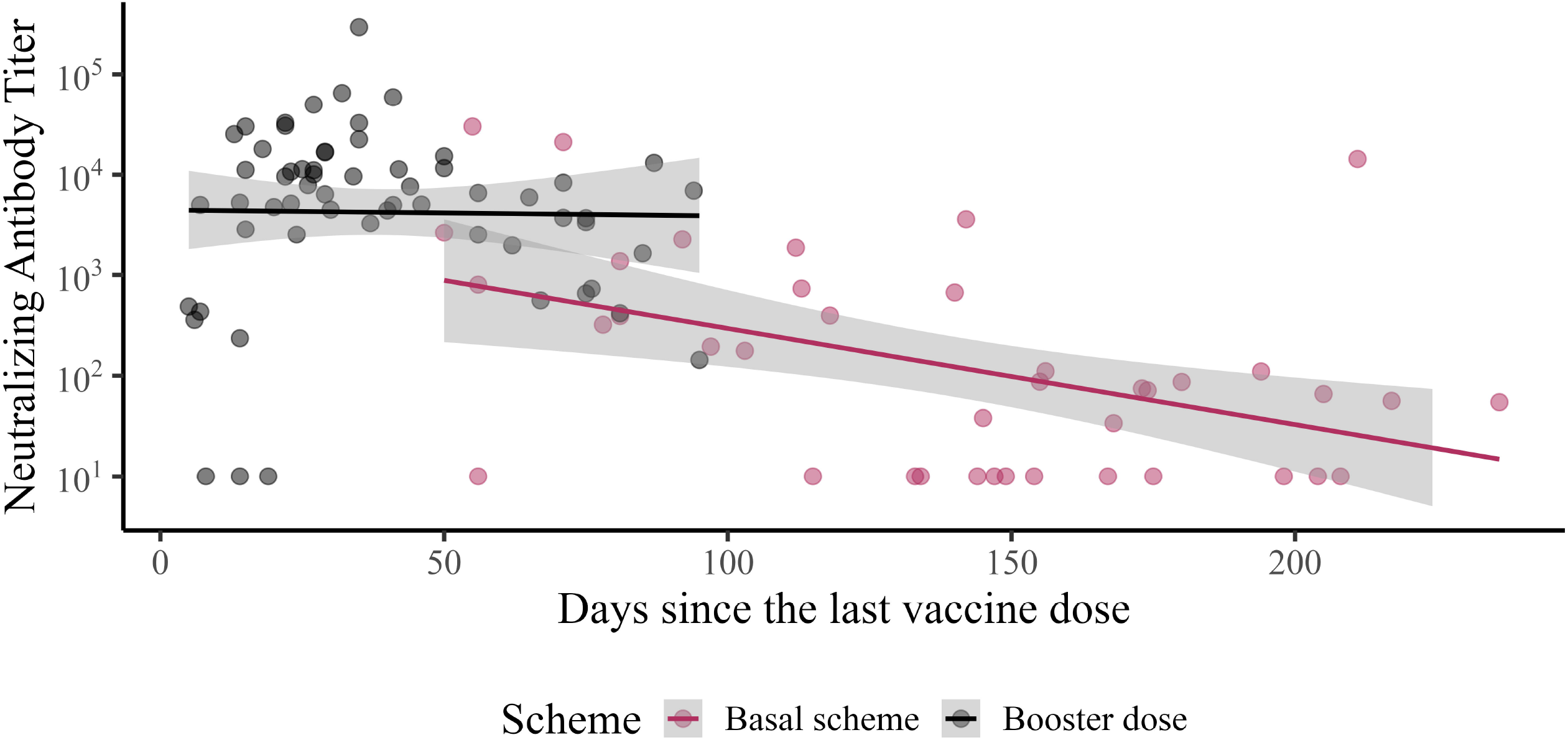

## Conclusions

Our results demonstrate that vaccination with a booster dose significantly improves the neutralization of the virus, and this effect may be associated with the relatively lower impact of the circulation of the Delta variant observed in Chile compared to the previous SARS-CoV-2 variants in terms of cases, hospitalizations, and deaths (10). By December 2021, 84.1% of the Chilean population had received a basal scheme vaccination and 56.1% a booster dose (10). People with natural infection had a similar level of nAbs compared to people vaccinated with the basal schemes. However, nAbs levels in both groups, natural infection, and basal schemes, were significantly lower than those with booster doses, reinforcing the importance of universal vaccination, regardless of the history of the disease, as a strategy that confers higher protection. Likewise, our results demonstrate the higher immunogenic potency of the mRNA vaccines, both in the basal and the booster dose schemes (5,11,12). Other studies on healthcare workers from Chilean institutions support the higher neutralizing titers triggered by mRNA vaccines’ basal scheme (13). A possible explanation might be the loss of antigenic sites in inactivated vaccines which only are exposed on a pre-fusion architectonic state of Spike, which is necessary for infection dynamics (14).

Nevertheless, the heterologous booster scheme, combining inactivated and mRNA vaccines (CCP), displayed a heterogeneous response, including 15% of subjects without nAbs; this figure is zero in the other two booster schemes (CCO and PPP), and 10% in those with PP basal scheme. Interestingly, the CCP group is younger than the CCO group (average 44 vs. 69 years old respectively), similar to PPP (average 44 years old), because the Chilean Health Authority restricted the use of ChAdOx1-S recombinant vaccine to people older than 55 years. A possible explanation for the proportion of non-responders with the CCP scheme may be the short time elapsed since the last vaccination. In fact, in 2 of the three subjects without nAbs, the sample collection was before 14 days, and this occurred in just 1 of the 17 nAbs responders. The neutralization analysis detected significant differences according to vaccine technologies not seen in measuring total SARS-CoV-2 antibodies (8). This added value, provided by neutralization studies, allows a deeper understanding of the antibody response to vaccines and natural infection to guide the public health response to the pandemic. Despite high vaccination coverage, we are still susceptible to new variants with the ability to evade the immune response, as was observed with the circulation of Omicron.

Finally, we found a lower nAbs response in smokers than non-smokers, consistent with studies suggesting a more inadequate humoral response in smokers (15).

The strength of this study includes the analysis of different vaccine technologies. In addition, it is a sample of subjects that comes from a population study and not from specific groups of the population. As for weaknesses, the moderate number of samples analyzed by vaccine technology does not include neutralizing antibodies against different variants of SARS-CoV-2, such as Delta and Omicron. Although previous studies have shown a correlation in neutralization for the different variants of SARS-CoV-2 (13), relevant changes have been detected for those with a greater capacity to evade the immune response.

We conclude that the booster dose significantly improves the levels of neutralizing antibodies against SARS-CoV-2, regardless of the vaccination scheme or the levels acquired by natural infection. Also, mRNA vaccine technology is strongly associated with higher neutralizing antibody levels than inactivated virus vaccines.

## Data Availability

All data produced in the present study are available upon reasonable request to the authors

## Acknowledgments

This research has been supported by WHO Unity Studies, a global seroepidemiological standardization initiative, with funding to WHO by the COVID-19 Solidarity Response Fund and the German Federal Ministry of Health (BMG) COVID-19 Research and Development Fund.

We thank K. Chandran for provision of a seed stock of VSV-GFP-Spike SARS-CoV-2 pseudotype.

## Disclaimer

The opinions expressed by authors contributing to this journal do not necessarily reflect the opinions of the Centers for Diseases Control and Prevention or the Institutions with which the authors are affiliated.

## Biographical Sketch

Dr. Ximena Aguilera is full professor of Public Health and Director of the Center of Epidemiology and Health Policy at the Universidad del Desarrollo, Santiago, Chile. Previously, she was Senior advisor on Communicable Diseases in the Pan American Health Organization (PAHO) in Washington DC. Earlier, she was head of Health Planning Division and head of the Department of Epidemiology for the Chilean Ministry of Health. Her interests include applied epidemiology to infectious diseases, global health, and health systems.

## References

1. Magazine N, Zhang T, Wu Y, McGee MC, Veggiani G, Huang W. Mutations and Evolution of the SARS-CoV-2 Spike Protein. Viruses. 2022 Mar 19;14(3):640.

2. Muena NAA, García-Salum T, Pardo-Roa C, Jos E Avenda∼ M, Serrano EF, Levican J, et al. Induction of SARS-CoV-2 neutralizing antibodies by CoronaVac and BNT162b2 vaccines in naïve and previously infected individuals. 2022; Available from: https://doi.org/10.1016/j.

3. Aguilera X, Mundt AP, Araos R, Weitzel T. The story behind Chile’s rapid rollout of COVID-19 vaccination. Travel Med Infect Dis. 2021;42(May):5–7.

4. Khoury DS, Cromer D, Reynaldi A, Schlub TE, Wheatley AK, Juno JA, et al. Neutralizing antibody levels are highly predictive of immune protection from symptomatic SARS-CoV-2 infection. Nat Med [Internet]. 2021;27(7):1205–11. Available from: http://dx.doi.org/10.1038/s41591-021-01377-8

5. Earle KA, Ambrosino DM, Fiore-Gartland A, Goldblatt D, Gilbert PB, Siber GR, et al. Evidence for antibody as a protective correlate for COVID-19 vaccines. Vaccine [Internet]. 2021;39(32):4423–8. Available from: https://doi.org/10.1016/j.vaccine.2021.05.063

6. Dieterle ME, Haslwanter D, Bortz RH, Wirchnianski AS, Lasso G, Vergnolle O, et al. A Replication-Competent Vesicular Stomatitis Virus for Studies of SARS-CoV-2 Spike-Mediated Cell Entry and Its Inhibition. Cell Host Microbe [Internet]. 2020;28(3):486–496.e6. Available from: https://doi.org/10.1016/j.chom.2020.06.020

7. Vial P, González C, Icaza G, Ramirez-Santana M, Quezada-Gaete R, Núñez-Franz L, et al. Seroprevalence, spatial distribution, and social determinants of SARS-CoV-2 in three urban centers of Chile. BMC Infect Dis. 2022;22(1).

8. Aguilera X, Gonzalez C, Apablaza M, Rubilar P, Icaza G, Ramirez-Santana M, et al. Immunization and SARS-CoV-2 antibodies seroprevalence in a country with high vaccination coveragelJ: Lessons from Chile. Res Sq [Internet]. 2022; Available from: https://www.researchsquare.com/article/rs-1548211/v1

9. Bergeri I, Ware H, Subissi L, Nardone A, Lewis HC, Li Z, et al. Global epidemiology of SARS-CoV-2 infection: a systematic review and meta-analysis of standardized population-based seroprevalence studies, Jan 2020-Dec 2021. medRxiv [Internet].2022; Available from: https://www.medrxiv.org/content/10.1101/2021.12.14.21267791v2

10. Roser M, Ritchie H, Ortiz-Ospina E, Hasell J. Coronavirus Pandemic (COVID-19). Our World Data [Internet]. 2020 Mar 5 [cited 2021 Apr 26]; Available from: https://ourworldindata.org/coronavirus

11. Barda N, Dagan N, Cohen C, Hernán MA, Lipsitch M, Kohane IS, et al. Effectiveness of a third dose of the BNT162b2 mRNA COVID-19 vaccine for preventing severe outcomes in Israel: an observational study. Lancet. 2021;398(10316):2093–100.

12. Munro APS, Janani L, Cornelius V, Aley PK, Babbage G, Baxter D, et al. Safety and immunogenicity of seven COVID-19 vaccines as a third dose (booster) following two doses of ChAdOx1 nCov-19 or BNT162b2 in the UK (COV-BOOST): a blinded, multicentre, randomised, controlled, phase 2 trial. Lancet. 2021;398(10318):2258–76.

13. Acevedo ML, Gaete-Argel A, Alonso-Palomares L, de Oca MM, Bustamante A, Gaggero A, et al. Differential neutralizing antibody responses elicited by CoronaVac and BNT162b2 against SARS-CoV-2 Lambda in Chile. Nat Microbiol. 2022;7(4):524–9.

14. Liu C, Mendonça L, Yang Y, Gao Y, Shen C, Liu J, et al. The Architecture of Inactivated SARS-CoV-2 with Postfusion Spikes Revealed by Cryo-EM and Cryo-ET. Structure. 2020;28(11):1218–1224.e4.

15. Ferrara P, Gianfredi V, Tomaselli V, Polosa R. The Effect of Smoking on Humoral Response to COVID-19 Vaccines: A Systematic Review of Epidemiological Studies. Vaccines. 2022;10(2):1–16.

